# Age-Dependent Sex Differences in the Prevalence and Prognosis of Multimorbid Conditions in Individuals with Heart Failure in Japan

**DOI:** 10.1101/2025.05.27.25328463

**Authors:** Shuhei Tara, Yayoi Tetsuou Tsukada, Takuya Nishino, Katsuhito Kato, Takeshi Yamamoto, Mitsunori Maruyama, Eitaro Kodani, Nobuaki Kobayashi, Akihiro Shirakabe, Kuniya Asai

**Affiliations:** Department of Cardiovascular Medicine, Nippon Medical School, Tokyo, Japan; Department of General Medicine and Health Science, Nippon Medical School, Tokyo, Japan; Department of Health Policy and Management, Nippon Medical School, Tokyo, Japan; Department of Hygiene and Public Health, Nippon Medical School, Tokyo, Japan; Division of Cardiovascular Intensive Care, Nippon Medical School Hospital, Tokyo, Japan; Department of Cardiovascular Medicine, Nippon Medical School Musashi-Kosugi Hospital, Kanagawa, Japan; Department of Cardiovascular Medicine, Nippon Medical School Tama Nagayama Hospital, Tokyo, Japan; Department of Cardiovascular Medicine, Nippon Medical School Chiba Hokusoh Hospital, Chiba, Japan; Division of Intensive Care Unit, Nippon Medical School Chiba Hokusoh Hospital, Chiba, Japan

**Keywords:** cardiovascular disease, mortality, rehospitalization, gender difference

## Abstract

**Background:** Multimorbidity is common among patients with heart failure (HF) and contributes to poor prognosis; however, the influence of age group and sex differences on the prevalence and outcomes of multimorbidity remains unclear.

**Methods:** This multicenter retrospective study included 3,004 hospitalized patients with HF. Multimorbidity was defined as the presence of two or more comorbidities and was quantified for stratification of comorbidity burden using the age-adjusted Charlson Comorbidity Index (CCI). Patients were dichotomized into high-and low-CCI groups based on the median CCI and were evaluated for prognosis using a composite endpoint of all-cause death or HF readmission.

**Results:** Multimorbidity increased with age but declined slightly in individuals aged > 85 years. And sex differences were observed, with males demonstrating a steeper increase in multimorbidity prevalence than females. Event-free survival rates were lower in the high-CCI group (hazard ratio [HR], 1.786; 95% confidence interval [CI], 1.483–2.151), consistent across sexes (males: HR, 1.927; 95% CI, 1.520–2.443; females: HR, 1.581; 95% CI, 1.171–2.135).

Among individuals aged ≥75 years with a high CCI, males had a stronger association with events than females (HR, 1.334; 95% CI, 1.031–1.727).

**Conclusions:** In individuals with HF, sex differences were evident in the prevalence of multimorbidity by age group and were associated with prognosis in older populations with a high comorbidity burden. Recognizing these differences is essential for tailoring HF management strategies to improve outcomes in individuals with multimorbid HF.

**Clinical trial registration:** URL: https://www.umin.ac.jp/ctr; unique identifier: UMIN000054854

**Clinical perspective What Is New?:**

- This multicenter study demonstrated that age-dependent sex differences exist in both the prevalence and prognostic impact of multimorbidity among Japanese individuals with heart failure.
- A three-way interaction analysis (age × sex × comorbidity burden) utilizing restricted cubic splines revealed that, among individuals with a high comorbidity burden, sex-related prognostic differences became increasingly pronounced with age, with older males exhibiting a higher risk than females.

**What Are the Clinical Implications?:**

- Incorporating age-and sex-specific risk assessments based on comorbidity burden may enhance individualized management strategies and improve clinical outcomes in individuals with heart failure, particularly in older adults with multiple comorbidities.

## Introduction

In aging societies, one of the major challenges is providing adequate healthcare for individuals with multimorbidity, defined as the presence of two or more chronic conditions in an individual^1,2^. Over 70% of adults develop cardiovascular disease (CVD) by the age of 70 years, and more than two-thirds of these individuals also experience non-CVD comorbidities, such as diabetes, chronic kidney disease, or chronic obstructive pulmonary disease^3,4^. Consequently, multimorbidity is particularly prevalent among older adults with CVD and is associated with a higher risk of hospital readmission, reduced quality of life, and worse survival outcomes^5^.

Heart failure (HF) represents the terminal stage of many CVDs and is characterized by complex clinical manifestations. As a result, individuals with HF commonly exhibit multimorbid conditions, further worsening their prognosis^6^. In Japan, which has become a super-aging society, the increase in individuals with HF, referred to as a “heart failure pandemic”, has placed a significant strain on the healthcare delivery system and led to a rise in medical expenses^7,8^.

Treatment strategies increasingly emphasize the need to consider diverse individual characteristics. However, the understanding and application of patient-specific factors among individuals with HF remain limited. For example, older females with HF are more likely to present with preserved ejection fraction and comorbid conditions, such as anemia or depression, whereas males often exhibit reduced ejection fraction and higher rates of ischemic heart disease^9^. These differences are often underappreciated in clinical practice and may contribute to disparities in treatment outcomes.

Based on these backgrounds, we focused on the potential age group and sex differences in multimorbid conditions among individuals with HF, as these differences may influence clinical outcomes. Therefore, this study aimed to investigate the age group and sex differences in the prevalence of multimorbidity and the prognostic impact of comorbidity burden among individuals with HF using the age-adjusted Charlson Comorbidity Index (CCI), a scoring system that incorporates specific comorbidities to estimate prognosis^10^.

## Methods

### Data Availability

The deidentified participant data will not be shared.

### Study Design and Data Collection

This multicenter retrospective cohort study was conducted using an inpatient database and follow-up data from four affiliated hospitals of Nippon Medical School: Nippon Medical School Hospital, Musashi Kosugi Hospital, Tama Nagayama Hospital, and Chiba Hokusoh Hospital. The database was constructed using a combination of Diagnosis Procedure Combination (DPC) data and medical record data. This study was approved by the Central Ethics Review Committee of Nippon Medical School (approval number: M-2024-178) and was conducted in accordance with the Declaration of Helsinki. Participant consent was obtained using the opt-out method.

The study population consisted of individuals aged 18 years or older with acute HF who were urgently admitted to the Department of Cardiovascular Medicine or the Cardiovascular Intensive Care Unit of the four affiliated hospitals of Nippon Medical School between April 2018 and September 2023 (**Figure 1**). Individuals with acute HF were identified as those with an ICD-10 code (I50X) for HF, recorded as the condition requiring the most medical resources, the primary diagnosis at the time of hospitalization, or the condition leading to hospitalization. After excluding individuals who died during hospitalization, 3,004 individuals who survived until discharge were included in the analysis.

**Figure 1.** Study population and inclusion criteria.

The primary endpoint of this study was the composite outcome of all-cause death and readmission due to HF within 1 year of hospital discharge.

### Variables

The DPC captured data on all hospitalized individuals, including demographics (age, sex), physical metrics (height, weight, body mass index [BMI] [calculated by dividing body weight in kilograms by the square of height in meters]), and clinical details (comorbidities, procedures during hospitalization, and medications at discharge^11^). Ultrasound left ventricular ejection fraction (LVEF) and blood test data at discharge, defined as the most recent sampling within 14 days before discharge were obtained from medical records.

### Definition of Comorbidities in Individuals with HF

Chronic conditions were defined using the CCI^10^ and identified using ICD-10 codes^12–14^. In this study, the following 18 chronic conditions, excluding HF from the basic 19 conditions of the CCI, were selected to define comorbidity in individuals with HF (**Supplementary Table 1)**: acquired immunodeficiency syndrome/human immunodeficiency virus, cerebrovascular disease, chronic pulmonary disease, dementia, diabetes with chronic complications, diabetes without chronic complications, hemiplegia or paraplegia, metastatic solid tumor, mild liver disease, moderate or severe liver disease, myocardial infarction, peptic ulcer disease, peripheral vascular disease, renal disease, rheumatologic disease, leukemia, lymphoma, and solid tumor without metastasis.

For epidemiological evaluation, individuals were categorized based on the number of comorbidities (0, 1, or ≥2), and multimorbidity was defined as the presence of two or more comorbidities. The comorbidity burden was quantified using the age-adjusted CCI as described below. The CCI was calculated by summing the weights assigned to each comorbidity, and additional points were added based on age (e.g., 1 point for 50–59 years, 2 points for 60–69 years, and 3 points for 70–79 years^10^). Stratification of comorbidity burden was conducted by dividing the cohort into two groups using the median age-adjusted CCI: low CCI (n = 1,478, CCI <6) and high CCI (n = 1,506, CCI ≥6).

### Statistical Analysis

Categorical variables are presented as numbers and percentages and were compared using Chi-square test. Continuous variables are presented as median values with interquartile ranges and were analyzed using the Mann–Whitney U test. For survival analysis, the Kaplan–Meier method was used to estimate the cumulative incidence of outcomes. Hazard ratios (HRs) and 95% confidence intervals (CIs) were calculated using Cox proportional hazards modeling.

Subgroup analyses were performed to evaluate sex differences in the primary endpoints across subgroups. Variables included in the subgroup analysis were age (<75 years vs. ≥75 years), LVEF (<40% vs. ≥40%), number of medications (<10 vs. ≥10), and BMI (<18.5, 18.5–<25.0, and ≥25.0). The HR and 95% CI for male versus female were calculated within each subgroup. Interaction *p*-values were calculated to assess the significance of sex-related interactions across subcategories. These analyses were conducted separately for the low-and high-CCI groups. To further evaluate the modifying effect of comorbidity burden on the relationship between age and sex in predicting outcomes, a multivariate Cox proportional hazards model was constructed, incorporating a three-way interaction term: age × sex × CCI group (low vs. high). Age was modeled as a continuous variable using restricted cubic splines (RCS) with four knots of 5th, 35th, 65th, 95th percentiles to account for potential nonlinear associations. The analysis was stratified by CCI group, and sex-specific hazard trajectories across age were visualized based on predicted HRs with 95% CIs.

Statistical significance was defined as a two-sided *p*-value of < 0.05. All analyses were performed using R software version 4.2.2 Patched (R Foundation for Statistical Computing, Vienna, Austria).

## Results

### Epidemiological Assessment: Age and Sex Differences of Multimorbidity

The prevalence in each age group for the three categories classified according to the number of comorbidities among individuals with HF (i.e., 0, 1, and ≥2) is shown in **Figure 2A**. The proportion of individuals with two or more comorbidities, defined as multimorbidity, steadily increased with age, peaking in the 71-75-year age group. However, after 85 years of age, this proportion declined slightly.

**Figure 2.** Epidemiological assessments for multimorbidity in individuals with heart failure. Individuals were categorized into three groups based on the number of comorbidities (0, 1, or ≥2). (**A**) The prevalence of the three groups across different age groups. (**B**) Gender-specific differences in the number of individuals within each age group, separately analyzed for males and females.

When the prevalence of the three groups, classified by the number of comorbidities (i.e., 0, 1, and ≥2), was evaluated by age group and sex, sex differences were observed (**Figure 2B**). Among males, the number of individuals with two or more comorbidities consistently exceeded those without comorbidities across all age groups aged 51 years or older. This gap gradually widened with advancing age, peaking at approximately 81–85 years of age. In contrast, among females, the number of individuals with two or more comorbidities was only slightly higher than among those without comorbidities. However, this trend reversed in individuals aged 81 years or older, with the number without comorbidities surpassing those with two or more.

### Patient Characteristics by Comorbidity Burden

The characteristics of individuals with low CCI (CCI <6) and high CCI (CCI ≥6) are summarized in **Table 1**. Individuals with HF in the high CCI group were significantly older and included a higher proportion of females than those in the low CCI group. The number of comorbidities included in the CCI criteria was two [1–2] in the high CCI group and zero [0–1] in the low CCI group. Individuals in the high CCI group also had longer hospital stays and lower BMI. The proportion of individuals with HF and reduced LVEF was lower, whereas the proportion with preserved LVEF was higher in the high CCI group compared to the low CCI group. Additionally, individuals in the high CCI group were prescribed a greater number of medications at discharge. Significant differences in the use of various medications and laboratory parameters were observed between the two groups. The high CCI group exhibited a higher prevalence of multimorbidity, as detailed in **Table 2**.

**Table 1.** Clinical characteristics of patients.

| **Variables** | **Low CCI**<br>**(n=1490)** | **High CCI**<br>**(n=1514)** | **p**<br>**value** |
| --- | --- | --- | --- |
| Age, years | 73 [62–82] | 83 [77–88] | <0.001 |
| Age category, years |  |  | <0.001 |
| <51 | 155 (10.4) | 5 (0.3) |  |
| 51–60 | 191 (12.8) | 22 (1.5) |  |
| 61–70 | 307 (20.6) | 102 (6.7) |  |
| 71–80 | 423 (28.4) | 389 (25.7) |  |
| 80< | 414 (27.8) | 996 (65.8) |  |
| Male sex | 907 (60.9) | 858 (56.7) | 0.021 |
| Length of hospital stay, days | 18 [12–27] | 21 [14–31] | <0.001 |
| Body mass index, kg/m<sup>2</sup> | 23.3 [20.4–26.4] | 22.3 [20.0–24.8] | <0.001 |
| LVEF, % | 40 [28–60] | 45 [31–60] | <0.001 |
| LVEF category, % |  |  | <0.001 |
| <40 | 661 (46.5) | 544 (37.5) |  |
| 40–50 | 189 (13.3) | 268 (18.5) |  |
| 50< | 571 (40.2) | 637 (44.0) |  |
| Mechanical support devices |  |  |  |
| Mechanical respiratory supports | 412 (27.7) | 484 (32.0) | 0.266 |
| Mechanical Circulatory Support | 54 (3.6) | 43 (2.8) | <0.001 |
| Hemodialysis | 20 (1.3) | 97 (6.4) | <0.001 |
| Number of comorbidities | 0 [0–1] | 2 [1–2] | <0.001 |
| Number of medications | 9 [7–12] | 11 [8–13] | <0.001 |
| Medications at discharge |  |  | <0.001 |
| Beta blocker | 1137 (76.3) | 1035 (68.4) | 0.058 |
| ACE inhibitor/ARB | 822 (55.2) | 782 (51.7) | <0.001 |
| ARNI | 243 (16.3) | 157 (10.4) | <0.001 |
| SGLT2 inhibitor | 348 (23.4) | 256 (16.9) | <0.001 |
| MRA | 889 (59.7) | 706 (46.6) | <0.001 |
| Aspirin | 251 (16.8) | 418 (27.6) | <0.001 |
| P2Y12 inhibitor | 220 (14.8) | 405 (26.8) | 0.002 |
| Direct oral anticoagulant | 683 (45.8) | 607 (40.1) | 0.099 |
| Warfarin | 111 (7.4) | 139 (9.2) | 0.106 |
| Loop diuretic | 1119 (75.1) | 1176 (77.7) | 0.008 |
| Tolvaptan | 354 (23.8) | 425 (28.1) | <0.001 |
| Statin | 562 (37.7) | 692 (45.7) | 0.011 |
| Laboratory data at discharge |  |  |  |
| White blood cells, $\times 10^3/\text{mm}^3$ | 5.9 [4.7–7.2] | 5.8 [4.6–7.1] | <0.001 |
| Hemoglobin, g/dL | 12.6 [10.9–14.4] | 11.1 [9.9–12.6] | 0.191 |
| Platelet, $\times 10^4/\text{mm}^3$ | 221 [177–278] | 213 [169–268] | <0.001 |
| Albumin, g/dL | 3.5 [3.2–3.8] | 3.3 [2.9–3.6] | <0.001 |
| Total bilirubin, mg/dL | 0.60 [0.45–0.88] | 0.50 [0.40–0.72] | 0.769 |
| Blood urea nitrogen, mg/dL | 21 [16–29] | 26 [19–37] | <0.001 |
| eGFR, mL/min/1.73 $\text{m}^2$ | 51 [37–65] | 40 [26–54] | <0.001 |
| Uric acid, mg/dL | 6.4 [5.2–7.8] | 6.5 [5.1–7.8] | 0.134 |
| NT-proBNP, pg/mL | 1438 [609–3228] | 2517 [1042–5432] | 0.011 |
| Sodium, mEq/L | 140 [138–142] | 140 [137–142] | 0.001 |
| Potassium, mEq/L | 4.30 [4.00–4.60] | 4.30 [3.90–4.60] | 0.191 |

Categorical variables are presented as n (%). Continuous variables are presented as median (interquartile range). LVEF, left ventricular ejection fraction; ACE, angiotensin-converting enzyme; ARB, angiotensin II receptor blocker; ARNI, angiotensin receptor–neprilysin inhibitor; SGLT2, sodium-glucose cotransporter 2; MRA, mineralocorticoid receptor antagonist; eGFR, estimated glomerular filtration rate; NT-proBNP, N-terminal pro–B-type natriuretic peptide.

**Table 2.** Prevalence of comorbidity.

| **Comorbidity conditions** | **Low CCI (n=1490)** | **High CCI (n=1514)** |
| --- | --- | --- |
| AIDS / HIV | 0 (0.0) | 2 (0.1) |
| Cerebrovascular disease | 42 (2.8) | 171 (11.3) |
| Chronic pulmonary disease | 82 (5.5) | 251 (16.6) |
| Dementia | 12 (0.8) | 169 (11.2) |
| Diabetes with chronic complication | 54 (3.6) | 246 (16.2) |
| Diabetes without chronic complication | 263 (17.7) | 499 (33.0) |
| Hemiplegia or paraplegia | 1 (0.1) | 8 (0.5) |
| Metastatic solid tumor | 0 (0.0) | 14 (0.9) |
| Mild liver disease | 19 (1.3) | 72 (4.8) |
| Moderate or severe liver disease | 0 (0.0) | 1 (0.1) |
| Myocardial infarction | 108 (7.2) | 385 (25.4) |
| Peptic ulcer disease | 142 (9.5) | 379 (25.0) |
| Peripheral vascular disease | 23 (1.5) | 135 (8.9) |
| Renal disease | 53 (3.6) | 453 (29.9) |
| Rheumatology disease | 14 (0.9) | 33 (2.2) |
| Leukemia | 2 (0.1) | 11 (0.7) |
| Lymphoma | 0 (0.0) | 11 (0.7) |
| Malignant tumor | 5 (0.3) | 110 (7.3) |

Variables are presented as n (%). CCI, Charlson Comorbidity Index; AIDS, acquired immunodeficiency syndrome; HIV, human immunodeficiency virus.

### Sex Differences in High Comorbidity Burden

Sex differences in individual characteristics were evaluated among those with a high comorbidity burden (CCI ≥6). Females, compared with males, were older, had a higher prevalence of HF with preserved LVEF, a lower rate of hemodialysis, and were prescribed fewer medications, particularly sodium-glucose cotransporter 2 inhibitors and antiplatelets (**Supplementary Table 2**).

Among individuals with a high comorbidity burden (CCI ≥6), sex differences were also observed in comorbidity prevalence. Females had a higher prevalence of dementia and rheumatologic diseases and a lower prevalence of chronic pulmonary disease, diabetes with chronic complications, myocardial infarction, and renal disease compared with males (**Supplementary Table 3**).

### Comorbidity Burden and Survival Analysis

In the overall population, 524 events occurred during the 1-year follow-up period, including 126 all-cause deaths and 398 HF readmissions. The event-free survival rate was significantly lower in the high CCI group compared with the low CCI group (low CCI group: 82.7% vs. high CCI group: 70.9%; *p* <0.001; HR: 1.786 [95% CI: 1.483–2.151]; **Figure 3A**).

**Figure 3.** Survival analyses for primary endpoints at 1-year post-discharge. Individuals were classified into two groups based on the Charlson Comorbidity Index (CCI): the low CCI group (CCI <6) and the high CCI group (CCI ≥6). The cumulative incidence of outcomes was compared between the groups in (**A**) the overall cohort, (**B**) males, and (**C**) females. HF, heart failure; HR, hazard ratio.

Similar trends were observed when stratified by sex. Among males, the event-free survival rate was significantly lower in the high CCI group (low CCI group: 83.1% vs. high CCI group: 69.8%; *p* <0.001; HR: 1.927 [95% CI: 1.520–2.443]; **Figure 3B**). Among females, the event-free survival rate was also lower in the high CCI group (low CCI group: 82.1% vs. high CCI group: 72.7%; *p* =0.003; HR: 1.581 [95% CI: 1.171–2.135]; **Figure 3C**). Although the HR was slightly lower in females than males, the association between higher comorbidity burden and poorer outcomes remained significant.

Subgroup analysis demonstrated that, among individuals with a low CCI (CCI <6), no significant association between sex and the primary endpoints was observed across any subgroup, and the interaction *p*-values similarly indicated no significant interactions between sex and these factors (**Figure 4A**). Conversely, among individuals with a high CCI (CCI ≥6), males aged ≥75 years exhibited a significantly higher association with the primary endpoints compared with females (HR: 1.334, 95% CI: 1.031–1.727), whereas no significant difference was observed among individuals aged <75 years (HR: 0.679, 95% CI: 0.346–1.331) (**Figure 4B**). The interaction *p*-value for age was 0.067, suggesting a potential modifying effect of sex in this subgroup, although statistical significance was not reached. In other subgroups, including those based on LVEF, number of medications, and BMI, no significant associations between sex and the primary endpoints were observed in any category, and the interaction *p*-values indicated no significant interactions between sex and these factors.

**Figure 4.** Subgroup analysis to assess sex differences for primary endpoints. HRs for males versus females were calculated within each subgroup. Interaction *p*-values were also calculated to assess the significance of sex-related interactions across the categories. These analyses were performed for (**A**) the low CCI group (CCI <6) and (**B**) the high CCI group (CCI ≥6). In addition, (**C**) a three-way interaction analysis (age × sex × CCI group) was conducted using Cox proportional hazards models with restricted cubic splines for age to examine how comorbidity burden modifies the age-and sex-related prognostic differences. CCI, Charlson Comorbidity Index; HR, hazard ratio; CI, confidence interval; BMI, body mass index (kg/m²).

In the three-way interaction model incorporating RCS for age, predicted risk showed a nonlinear association with age, with distinct trajectories stratified by comorbidity burden and sex (**Figure 4C**). Among individuals in the low CCI group, the HR increased sharply with advancing age, particularly among males, whereas the increase was more gradual among females. In contrast, in the high CCI group, the predicted risk curves for males and females intersected around the age of 70 years, indicating that sex-related differences in prognosis may shift direction with increasing age.

Cox regression analysis was performed to evaluate prognostic factors among individuals stratified by comorbidity burden (**Table 3**). In the multivariable analysis, age ≥75 years independently associated with worse outcomes in both groups (low CCI: HR: 2.050, 95% CI: 1.443–2.910, p<0.001; high CCI: HR: 1.580, 95% CI: 1.112–2.246, p = 0.011). BMI ≥25 was also associated with better prognosis (low CCI: HR: 0.614, 95% CI: 0.424–0.888, p = 0.010; high CCI: HR: 0.649, 95% CI: 0.465–0.905, p = 0.011). Use of ≥10 medications was a significant predictor of worse outcomes only in the low CCI group (HR: 1.646, 95% CI: 1.197– 2.264, p = 0.002), whereas BMI <18.5 kg/m^2^ was significantly associated with poor prognosis only in the high CCI group (HR: 1.863, 95% CI: 1.363–2.547, p<0.001). Male sex was independently associated with worse outcomes in the low CCI group (HR: 1.460, 95% CI: 1.042–2.046, p = 0.028) but not in the high CCI group.

**Table 3.** Cox regression analysis Low CCI.

| **Multivariable analysis** |  |  |  |
| --- | --- | --- | --- |
| **Variables** | **HR** | **95% CI** | **p value** |
| Age $\geq 75$ | 2.050 | 1.443–2.910 | <0.001 |
| Male | 1.460 | 1.042–2.046 | 0.028 |
| LVEF <40% | 0.821 | 0.587–1.150 | 0.252 |
| Number of medications $\geq 10$ | 1.646 | 1.197–2.264 | 0.002 |
| BMI <18.5 kg/m<sup>2</sup> | 1.420 | 0.905–2.227 | 0.127 |
| BMI $\geq 18.5$ kg/m<sup>2</sup>, <25 kg/m<sup>2</sup> | | | |
| BMI $\geq 25$ kg/m<sup>2</sup> | 0.614 | 0.424–0.888 | 0.010 |

**High CCI**
| **Multivariable analysis** |  |  |  |
| --- | --- | --- | --- |
| **Variables** | **HR** | **95% CI** | **p value** |
| Age $\geq 75$ | 1.580 | 1.112–2.246 | 0.011 |
| Male | 1.220 | 0.941–1.582 | 0.134 |
| LVEF <40% | 0.973 | 0.753–1.258 | 0.837 |
| Number of medications $\geq 10$ | 1.165 | 0.897–1.513 | 0.252 |
| BMI <18.5 kg/m<sup>2</sup> | 1.863 | 1.363–2.547 | <0.001 |
| BMI $\geq 18.5$ kg/m<sup>2</sup>, <25 kg/m<sup>2</sup> | | | |
| BMI $\geq 25$ kg/m<sup>2</sup> | 0.649 | 0.465–0.905 | 0.011 |

CCI, Charlson comorbidity index; HR, hazard ratio; CI, confidence interval; LVEF, left ventricular ejection fraction; BMI, body mass index

## Discussion

This study demonstrates that sex differences exist in the prevalence of multimorbidity among patients with HF across different age groups. Higher comorbidity burden was consistently associated with worse outcomes, including all-cause death and HF readmission, in both sexes. Notably, older males with a high comorbidity burden were more strongly associated with adverse outcomes than their female counterparts. Japan is one of the world’s most rapidly aging societies. Patients with HF in Japan are aging, with the proportion of females also increasing^7^. Comparative studies have shown that Japanese patients with HF tend to be older and are more often female than patients in other Asian or European populations^7,15,16^. Although this study was conducted solely in Japan, its findings may offer valuable insights for other countries facing similar demographic transitions. However, differences in healthcare systems, socioeconomic contexts, and disease management practices may limit the generalizability of the results. Future international comparative studies are warranted to validate whether the observed sex-specific patterns of multimorbidity and HF outcomes observed here also apply to patients in other healthcare systems.

Our findings underscore the importance of adopting a comprehensive management approach that incorporates multimorbidity into HF care. Multimorbidity in HF can be classified into two categories: concordant conditions (e.g., HF and hypertension), which share pathophysiological pathways and treatment strategies, and discordant conditions (e.g., HF and osteoarthritis), which have unrelated mechanisms but still impact overall care^5,17^. Despite the need to address both types, current disease-specific guidelines often lack recommendations for managing discordant comorbidities in patients with HF. This gap may be attributed to the complexity of individualized treatment planning and the lack of evidence supporting integrated care approaches^5^. Furthermore, the observed sex differences in multimorbid conditions underscore the importance of incorporating sex-specific considerations into clinical guidelines and treatment plans^18^. However, this study did not identify specific factors related to multimorbidity, such as biological and clinical features, lifestyle and health behaviors, or social and mental factors, that contribute to sex differences^19^. Additional research is warranted to identify the underlying mechanisms and develop more personalized therapeutic strategies.

To quantify comorbidity burden, we used the age-adjusted Charlson Comorbidity Index^6,20^. Previous research using Japanese nationwide claims data demonstrated a strong association between higher CCI and poor outcomes^12^. Furthermore, non-cardiac comorbidities are known to exacerbate HF progression, with an increasing number of comorbidities associated with higher hospitalization rates^21^. However, the prognostic implications of comorbidities may vary depending on the type and severity of the condition, as well as patient age. Thus, quantifying both comorbidity burden and age is essential for accurate risk stratification, especially among elderly populations in rapidly aging societies. In our cohort, patients with a CCI ≥6 were classified as having a high comorbidity burden. This threshold is comparable to previous findings suggesting that higher age-adjusted CCI scores predict adverse outcomes in HF^6,22^. Minor discrepancies may be attributed to variations in population demographics, regional healthcare practices, or operational definitions of comorbidities.

In the epidemiological assessment of this study, the proportion of patients with multimorbidity steadily increased with age, peaking in the 75–84-year age group and slightly declining after 85 years. This pattern suggests that while multimorbidity becomes increasingly common with advancing age, there may be a slight reduction among the oldest age group, possibly due to survival bias or other age-related factors. Significant sex-based differences in the distribution of multimorbidity were also observed, particularly in older age groups, with males showing a steeper increase in the prevalence of multiple comorbidities than females.

These findings may inform future prospective research focused on sex differences in multimorbidity among older populations.

Multivariate Cox regression analysis demonstrated that among patients with a high comorbidity burden, older age and low body weight, rather than sex—were predominantly associated with adverse outcomes. In contrast, male sex was independently associated with worse prognosis only in the low CCI group, suggesting that sex differences may be more evident in patients with a low comorbidity burden. The discrepancy between multivariable Cox analysis and subgroup analysis, where older males with a high comorbidity burden demonstrated a greater association with adverse outcomes than females, may reflect age-dependent modification of sex effects on prognosis, as confirmed by spline-based three-way interaction analysis. These results underscore the importance of incorporating age-and sex-specific considerations into risk stratification and management strategies for patients with HF and high comorbidity burden.

This study had some limitations. First, because this was a retrospective analysis, the findings are subject to potential selection bias. Second, reliance on ICD-10 coding may have led to underreporting or misclassification of comorbidities. Third, the lack of detailed data on treatment regimens and their interaction with comorbidities limits the interpretability of the prognostic outcomes. Future studies should use prospective designs to confirm causality and investigate the effect of integrated multimorbidity-focused interventions, ideally stratified by sex—on clinical outcomes in HF.

## Conclusions

In individuals with HF, sex differences in the prevalence and prognostic impact of multimorbidity varied with age. Among older individuals with a high comorbidity burden, males were at a higher risk of adverse outcomes compared with females. Recognizing and addressing these age-and sex-specific disparities is essential for improving management strategies and outcomes in individuals with multimorbidity and HF.

## Non-standard Abbreviations and Acronyms

ACE: angiotensin-converting enzyme
ARB: angiotensin II receptor blocker
ARNI: angiotensin receptor neprilysin inhibitor
CVD: cardiovascular disease
eGFR: estimated glomerular filtration rate
ICD-10: International Classification of Diseases 10th Revision
LVEF: left ventricular ejection fraction
NT-proBNP: N-terminal pro-brain natriuretic peptide
SGLT2: sodium-glucose cotransporter 2

## Data Availability

The datasets generated during and/or analysed during the current study are available from the corresponding author on reasonable request.

## Acknowledgements

The authors extend their sincere thanks to all individuals involved in patient care, including emergency staff, technicians, medical engineers, nurses, pharmacists, physicians, and surgeons at Nippon Medical School Hospital, Musashi-Kosugi Hospital, Tama Nagayama Hospital, Chiba Hokusoh Hospital, and Nippon Medical School.

## Supplemental Material

Supplementary Table 1: Conditions of the Charlson Comorbidity Index

Supplementary Table 2: Sex differences in the clinical characteristics of patients with heart failure and a high-comorbid burden

Supplementary Table 3: Sex difference in the prevalence of comorbidity in high comorbid burden

